# Rise and Regional Variations in Schedule II Stimulant Use in the United States

**DOI:** 10.1101/2020.04.28.20069054

**Authors:** Sneha M. Vaddadi, Nicholas J. Czelatka, Belsy D. Gutierrez, Bhumika C. Maddineni, Carlos D. Torres-Teran, Daniel N. Tron, Kenneth L. McCall, Brian J. Piper

## Abstract

**Objective:** There is a need to better understand recent trends in stimulant usage. This report compares the pharmacoepidemiology of three Schedule II stimulants in the United States from 2010 – 2017.

**Methods:** Drug weights were extracted from the Automated Reports and Consolidated Ordering Systems (ARCOS) for amphetamine, methylphenidate, and lisdexamfetamine. Total grams per drug were averaged across all states and compared from 2010–2017. Median stimulant daily dosage per patient user was determined from electronic medical records for a regional analysis.

**Results:** There was a rise in amphetamine (+67.5%) and lisdexamfetamine (+76.7%) use from 2010–2017. The change in methylphenidate (−3.0%) was modest. Regional analysis indicated that persons/day usage of stimulants in the west was lower than that of other US regions from 2014–2017. There was a negative correlation (*r*(48) = −0.43 to −0.65, *p* < .05) between the percent Hispanic population per state and the daily dose/population per stimulant.

**Conclusion:** The increasing amphetamine and lisdexamfetamine, but not methylphenidate, use may be explained by a rise in adult ADHD diagnoses and treatment. Regional analysis indicates that the use of stimulants in the west may be distinct from that in other regions. The lower stimulant use in areas with greater Hispanic population may reflect socioeconomic factors. Further research is needed on social factors impacting stimulant use and reasons for the pronounced regional variance.

## Introduction

Stimulants are used in treatment for Attention Deficit Hyperactivity Disorder (ADHD)- a disorder characterized by hyperactivity, inattention, executive function deficits and emotional dysregulation^1,2^. ADHD is one of the more common biopsychosocial disorders, with a growth of 3.4% per year from 1998 – 2008 and a prevalence of 11% in children in 2011^3^. Based on data from the National Survey of Children’s Health, the percent of medicated ADHD nationally in children age 4-17 rose from 4.8% to 6.1% from 2007–2011^3^. ADHD diagnosis and treatment extends beyond childhood. With a revised ADHD criterion released in 2013 in the DSM-5, ADHD diagnoses have been more inclusive of adolescents and adults. As of 2018, approximately 4% of US adults are afflicted with ADHD and two-thirds of children continue to experience at least one ADHD symptom throughout their lives^4^. Despite the growth in ADHD, there is variation within cultural communities as well. Despite being one of the largest ethnic minorities in the United States, Hispanic youths have less ADHD diagnoses and stimulant use^5,6^.

The common ADHD stimulants include amphetamine, lisdexamphetamine, and methylphenidate. Atomoxetine is a selective norepinephrine reuptake inhibitor used as a non-stimulant alternative for ADHD treatment^7^. Stimulant use has been associated with mild adverse effects such as appetite and sleep disturbances that impact quality of life, but the long-term adverse effects of these substances are not well established^8^. A meta-analysis of cross-sectional Positron Emission Tomography investigations showed that long-term blockade of the dopamine transporter caused neuroadaptive striatal elevations in this protein^9^. This finding was subsequently confirmed in a longitudinal report^10^. Patients prescribed stimulants had a nine-fold elevated risk of developing basal ganglia and cerebellar disorders^11^. Non-medical use of stimulants is also appreciable. The Monitoring the Future survey of recreational drug use determined that 4.6% of 12^th^ graders misused Adderall in 2018^12^.

With increasing prevalence of medicated ADHD in children and adults, there is a greater need to understand the extent of stimulant use nationally. This report utilized the US Drug Enforcement Administration’s Automated Reports and Consolidated Ordering Systems (ARCOS) comprehensive database to evaluate changes in use of amphetamine, methylphenidate, and lisdexamfetamine nationally from prior years to 2017. We extended upon past research by investigating the overall change of stimulant use from 2010–2017^13^. We then calculated daily dose values to investigate the change in use from 2016 and 2017. We also explored variations in use in the Hispanic population and geographical regions. Investigation of these databases will allow for a greater understanding of the most recent pattern of use in treatment of ADHD and recent fluctuations of use within the United States.

## Methods

### Data Sources

Stimulant data were extracted from the DEA’s Automated Reports and Consolidated Ordering Systems (ARCOS). This national database contains a yearly updated report of retail drug distribution by zip code and by state submitted by manufacturers and distributors to the DEA. Extracted data included total grams of stimulant use per drug per state from 2010 to 2017. Three Schedule II stimulants were examined: amphetamine, methylphenidate and lisdexamfetamine. Methamphetamine was not examined here due to low values. This database has been frequently used in prior pharmacoepidemiology reports^13,15^.

Our goal was to examine the change in stimulant use and change in number of patients utilizing Schedule II stimulants, but ARCOS data is limited to the total quantity of Schedule II stimulants distributed in a geographical location. To determine the median stimulant daily dosage per patient user, we used de-identified data from the electronic health record of Geisinger health system, an integrated health delivery system in central and northeastern Pennsylvania. With Geisinger EHR data from 2018 (n=88,202), we were able to estimate the median doses as 20 mg/day/person for methylphenidate and amphetamine and 40 mg/day/person for lisdexamfetamine. These values were used for daily dose calculations, which was used for a 2016–2017 and a regional comparison.

An analysis was completed with state-specific Hispanic population data. The percent Hispanic population per state was obtained from the demographic profiles from the Pew Research Center. Institutional Review Board approval was obtained from the University of New England and Geisinger.

### Data Analysis

Analysis by stimulant weight was completed by averaging state data per year for each stimulant and comparing use from 2010–2017. ARCOS aggregate data was divided by median values for daily dosage (mg/person/day) per stimulant that was determined from Geisinger EMR data. These values were named “Persons/day” and were used for a regional analysis by averaging state data within each region (Midwest, Northeast, South, West) for all three stimulants and compared from 2014–2017. Data was deemed to be significant (*p* < 0.05) after an unpaired t-test. A population-corrected analysis termed “Daily Dose/Population” was done by further dividing the ARCOS mg/daily dose by population per state for each drug. The percent change in these values per state per drug from 2016–2017 were calculated and compared. Values were deemed significant as 1.96 standard deviations above and below the mean. For all above calculations, outliers were determined through a Grubbs analysis and significant values were excluded. A heat map was constructed with Excel using the sum of ARCOS mg/daily dose value for all three stimulants divided by population for 2017. A similar calculation was done for 2016 and the percent change values were also depicted. A linear regression analysis was done with daily dose/population values and Hispanic population data per state in 2016 and 2017. Variance was reported as the SEM.

## Results

Stimulant use, by weight, increased 67.5% for amphetamine and 76.7% for lisdexamfetamine from 2010–2017 on average across all fifty states. In contrast, methylphenidate use decreased slightly (3.0%). For amphetamine and lisdexamfetamine, there was a significant increase in the total stimulant use compared to 2010 starting from 2014 (Figure 1A). Further investigation into the change from 2016–2017 was completed with a daily-dose and population-corrected analysis. The percent change in daily dose/population across fifty states (with New Mexico excluded as an outlier) was +4.6% for amphetamine, +2.3% for lisdexamfetamine, and −1.4% for methylphenidate (Figure 1B, C). The preponderance (85.0%) of states increased their amphetamine and over two-thirds (72.0%) increased their lisdexamfetamine use. In contrast, 86.0% of states decreased their methylphenidate use. Wisconsin, South Dakota and West Virginia were all significantly greater than the mean for amphetamine. Hawaii had a significantly greater value while Nevada and South Dakota had lower values compared to the national average for methylphenidate. For lisdexamfetamine, South Dakota had a significantly greater value while Wisconsin had a lower value relative to the mean (Figure 1D).

**Figure 1:**
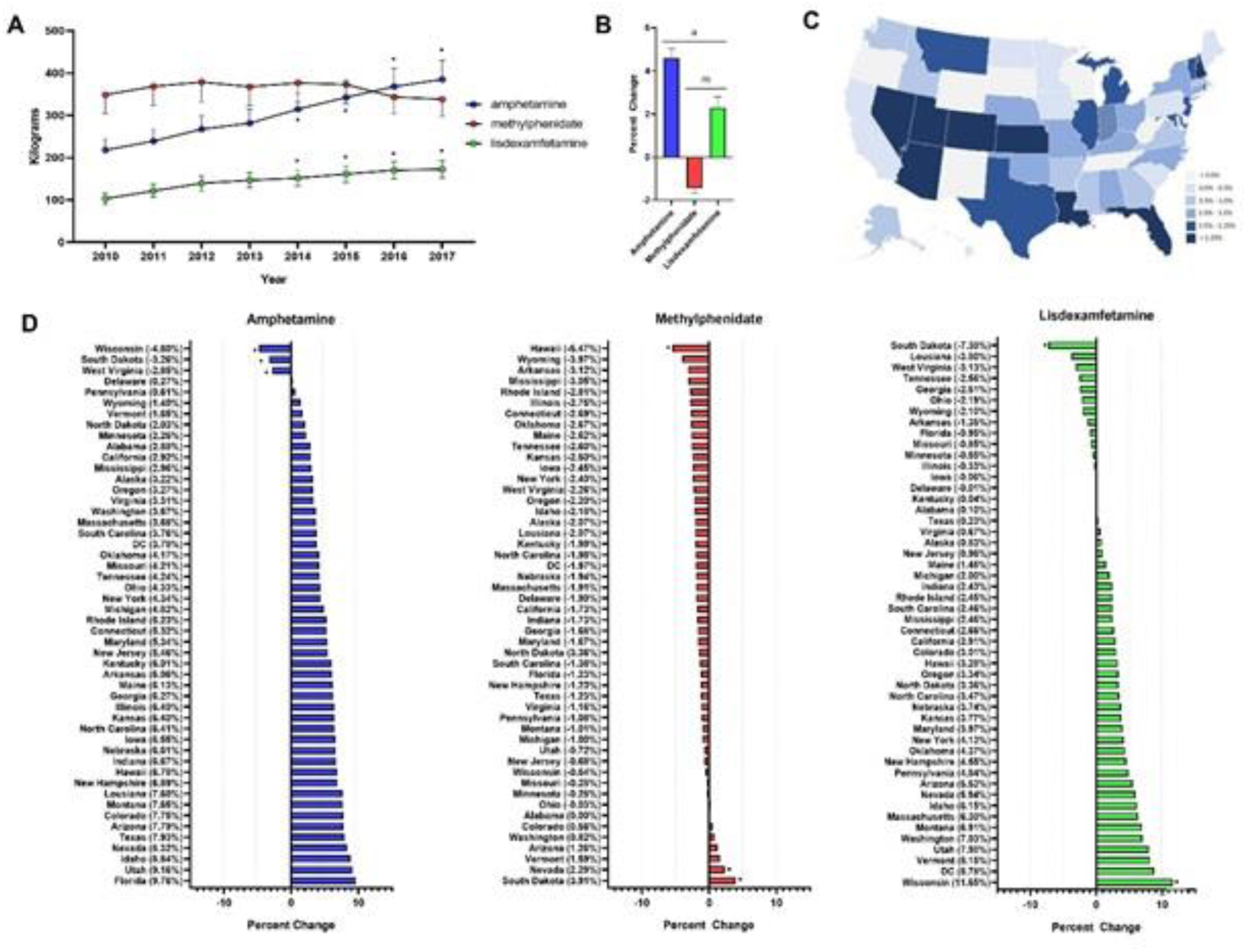
**A** Weight per stimulant per state showed a 67.5% and 76.7% increase in amphetamine and lisdexamfetamine (\**p* < 0.05 in comparison to 2010) and a 3% decrease in methylphenidate. **B** Average percent change of 50 states data in Daily Dose/Person from 2016–2017 for amphetamine (+4.6%), lisdexamfetamine (+2.3%), and methylphenidate (−1.4%). Lisdexamfetamine was significantly different from amphetamine (^a^*p* <0.05) and methylphenidate (^m^p < 0.05). **C** Heat map of United States depicting percent change in total stimulant daily dose/population per state from 2016–2017. **D** Percent change in daily dose and population adjusted analysis per state for amphetamine, methylphenidate, and lisdexamfetamine reveals 85.0% of states increased their amphetamine, 72.0% increased their lisdexamfetamine, and 86.0% states decreased their methylphenidate use. Significant states were marked if 1.96*SD greater or less than the mean for each stimulant. The percent change for New Mexico was excluded as an outlier in all graphs (−17.45% for amphetamine, −19.98% for methylphenidate, and −36.63% for lisdexamfetamine).

The daily dose/population values for 2017 are depicted in the heat map in Figure 2A, indicating pronounced regional variance. There was a six-fold difference between values for the highest (12.2) and lowest (2.9) states. Six states with the lowest values of daily dose/population were all in the Western region of the United States. A regional analysis was completed to further investigate geographic-based variability. A comparison of persons/day per region from 2014–2017 revealed a significant difference in the values for the Midwest, Northeast, and South compared to the West consistently from 2014–2017 (Figure 2B). Analysis with the percent Hispanic population and Daily dose/population identified a negative correlation for each stimulant in 2016. These findings were replicated for 2017 (Figure 3). States with a greater portion of Hispanic populations used less stimulants.

**Figure 2:**
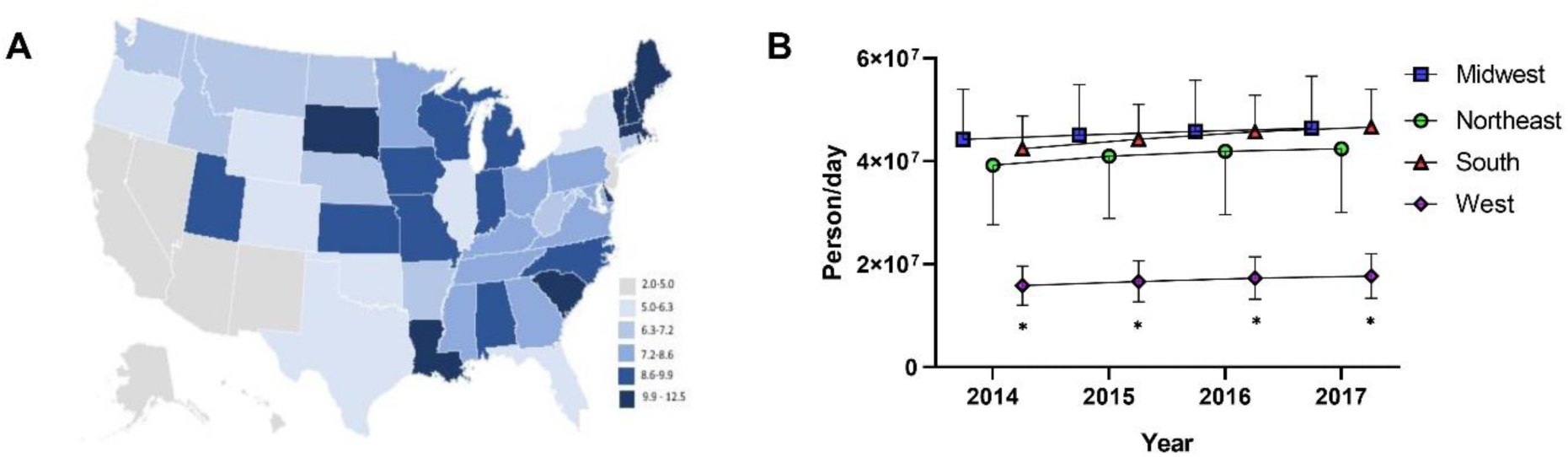
**A** Heat map for Daily Dose/Population per state for 2017 **B** Person/Day per Region from 2014–2017 indicated a significant difference with the West compared to the South, Midwest and Northeast from 2014–2017 (\**p* < 0.05). Time points were slightly offset for display purposes.

**Figure 3:**
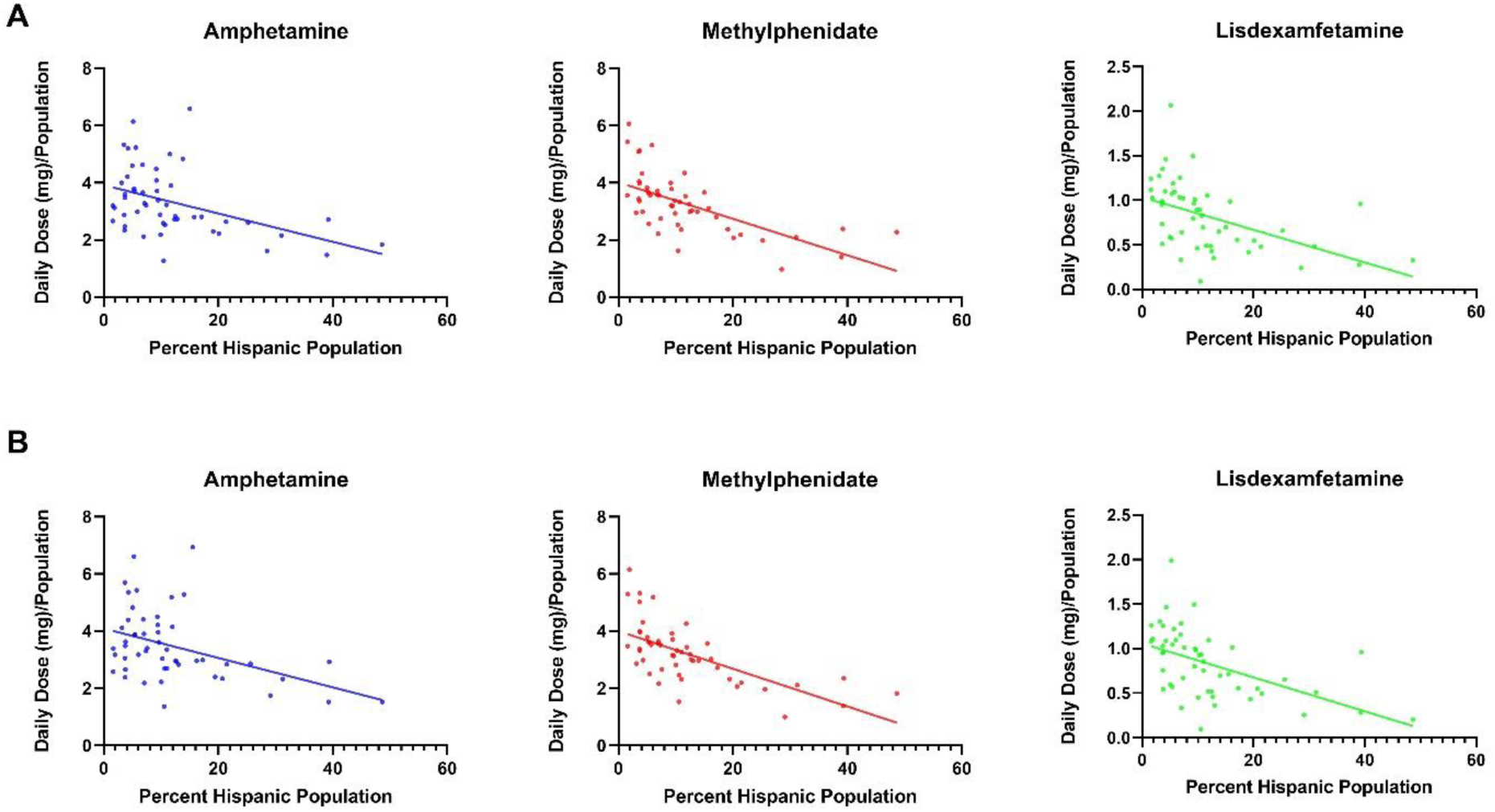
**A** Negative correlation between total Daily Dose/Population value and the percent Hispanic per state for 2016. (amphetamine: *r*(48) = −0.43, *p* < 0.01; methylphenidate: *r*(48) = −0.64, *p* < 0.0001; lisdexamfetamine: *r*(48) = −0.49, *p* ≤ 0.0001). **B** Negative correlations between total Daily Dose/Population value and percent Hispanic per state for 2017 (amphetamine *r*(48) = −0.43, *p* < 0.01; methylphenidate: *r*(48) = −0.65, *p* < 0.0001; lisdexamfetamine *r*(48) = −0.52, *p* ≤ 0.0001).

## Discussion

This study investigated trends in stimulant use in the United States in 2017 relative to prior years. Our data was indicative of an overall increase of Schedule II stimulant use, which is consistent with accounts of rising number of ADHD diagnoses^1^. An analysis for total grams from 2010–2017 and a daily dose and population-considered analysis from 2016–2017 indicated a rise in amphetamine and lisdexamfetamine and no appreciable change in methylphenidate.

This pattern of stimulant use change may be explained by a rise in adult ADHD diagnoses and treatment. The revision of ADHD criteria in the DSM-5 is more inclusive of adult ADHD and has led to adults meeting more requirements for diagnosis than that for the DSM-IV^17^. A 2018 study looking at ADHD treatment in privately-insured women aged 15-44 found a similar pattern in these three stimulants as this study, with the largest change in stimulant use being in the age range of 25-29 years^16^. Studies have also indicated variance in drug efficacy for long term treatment in adult ADHD. While methylphenidate is considered a first-line treatment for child and adolescent ADHD, the long-term (> 12 months) efficacy of stimulants is not well established^18^. There are also drug-drug interactions with many medications, including MAO oxidase inhibitors, vasopressors, and coumadin anticoagulants^2,24^. Lisdexamfetamine and mixed amphetamine salts were found to cause a significant improvement in adult ADHD symptoms without symptoms rebound after ceasing medication^19,25^. Alternate uses for stimulants outside of AHDH treatment may also contribute to these patterns in stimulant use. Lisdexamfetamine is a well-tolerated treatment for moderate to severe binge eating disorder^20^. Literature reviews have also explored the role of amphetamine and methylphenidate in treatment of apathy in Alzheimer’s patients and other neuropsychiatric conditions in the elderly^21^. Further exploration is needed on how the expansion of stimulant use in other neuropsychiatric conditions or obesity impacts usage trends.

The percent Hispanic population had a negative correlation with stimulant use per state for 2016 and 2017. Other studies have also indicated a lower stimulant use by Hispanic children compared to their non-Hispanic peers^14^. Young Hispanic adults and children have a significantly lower use of outpatient mental health services for mental health and substance abuse care^22^. The low rate of stimulant use among the Hispanic communities may be due to difficulties with access to healthcare. Prior to the implementation to the ACA in 2014, 30% of Hispanics reported no health insurance compared to 11% of non-Hispanic whites^6^. Along with social factors such as language barriers, cultural factors such as a perceived difference in the need for outpatient mental health care may also explain differences in resource utilization^23^. The cumulative effect of these factors may lead to individuals being unable or hesitant to seek medical attention for ADHD symptoms.

Finally, our regional analysis with data controlled for daily dosage found that the West has a significantly lower schedule II stimulant use compared to the South, Northeast, and Midwest. This pattern was seen in other studies spanning from 1998–2018 focusing on both child and adult ADHD, with the West having the lowest ADHD prevalence or change in stimulant use^13,26,27^. Though widely noted, this pattern has little explanation and may be due to various factors. A 2015 report suggests that with many states of higher altitude located primarily in the West, the altitude may serve as a protective factor against ADHD by increasing dopamine levels^26^. Cultural diversity may also play a role. In California, almost 40% of youths are Hispanic, an ethnicity that has significantly lower stimulant use^5^.

In conclusion, this report identified increases in use in amphetamine and lisdexamfetamine in the United States. Further investigation is needed to better understand the sociocultural or economic factors mediating the pronounced regional variance observed.

## Data Availability

Raw data is available from the Drug Enforcement Administration.

https://www.deadiversion.usdoj.gov/arcos/retail_drug_summary/index.html

## Acknowledgements

The generation of heat maps was done with the help of Daniel Kaufman. This project was also completed with the technical assistance of Iris Johnson.

